# *TMEM106B* analysis in diverse cohorts suggests no association with Parkinson’s disease risk

**DOI:** 10.64898/2026.09.28.26363833

**Authors:** Henry Mauricio Chaparro-Solano, Sajanth Kanagasingam, Ziv Gan-Or, Konstantin Senkevich, the Global Parkinson’s Genetic Program (GP2)

**Affiliations:** Facultad de Ciencias de la Salud, Universidad Autónoma de Bucaramanga – UNAB, Avenida 42 No. 48-11, Bucaramanga, 680003, Colombia; Genomic Sciences and Systems Biology, Cleveland Clinic Research, 9500 Euclid Avenue, Cleveland Clinic, Cleveland, Ohio, 44195, USA; Department of Molecular Medicine, Cleveland Clinic Lerner College of Medicine, Case Western Reserve University, Cleveland, Ohio, 44195, USA; Department of Human Genetics, McGill University, Montreal, Quebec, Canada; The Neuro (Montreal Neurological Institute-Hospital), McGill University, Montreal, Quebec, Canada; Department of Neurology and Neurosurgery, McGill University, Montréal, Quebec, Canada, Canada; Department of Specialized Medicine, Division of Medical Genetics, McGill University Health Centre, Montreal, Quebec, Canada

## Abstract

Recent evidence suggests that TMEM106B expression is reduced in the brains of patients with Parkinson’s disease (PD). However, an association between rare genetic variants in TMEM106B and PD risk has not been previously established. In this work, we investigated the association between rare TMEM106B variants and PD risk. We leveraged whole-genome sequencing data from 22,010 cases and 40,550 controls across diverse ancestries from the Global Parkinson’s Genetics Program (GP2), the Accelerating Medicines Partnership in Parkinson’s Disease (AMP-PD), and the UK Biobank (UKBB). A total of 4,581 unique rare variants were identified across all groups. We found no evidence that rare coding variants in TMEM106B were associated with PD risk, either individually or through cumulative burden. These findings suggest that rare variation in TMEM106B does not play a major role in PD susceptibility across the populations studied.

---

We read with great interest the recently published article by Meng and colleagues, entitled “TMEM106B deficiency exacerbates α-synuclein aggregation in Parkinson’s disease”. In this work, the authors present evidence showing that TMEM106B expression is reduced in the brains of patients with Parkinson’s disease. In addition, using animal and in vitro models, they demonstrate that knockdown of this gene leads to increased α-synuclein aggregation and delayed degradation. Altogether, these findings suggest a relevant association between TMEM106B function and Parkinson’s disease pathophysiology (1).

The *TMEM106B* gene (OMIM *613413) encodes the transmembrane protein 106B, which plays a role in endosome-lysosome morphology and acidification (2). As discussed by Meng et al., this may explain the biological association between *TMEM106B* and Parkinson’s disease risk, given that endosome-lysosome dysfunction is recognized as an important pathway in Parkinson’s disease pathophysiology (1).

*TMEM106B* has been implicated in multiple neurodegenerative conditions. Common variants in this gene were first identified as genetic risk factors for frontotemporal lobar degeneration with TDP-43 inclusions through a genome-wide association study (GWAS), which identified the minor allele of the lead single nucleotide polymorphism (SNP) rs1990622 as protective (4). Additional studies have suggested that the protective effect of *TMEM106B* is particularly pronounced in individuals who also carry *GRN* variants (4,5).

Notably, a direct genetic association between *TMEM106B* and Parkinson’s disease risk has not been previously reported, and *TMEM106B* has not been identified as a genome-wide significant locus in large-scale Parkinson’s disease GWAS (Figure 1) (6). One study reported that common *TMEM106B* variants may act as genetic modifiers of cognitive decline in Parkinson’s disease (7). However, this finding has not yet been independently replicated. These observations prompted us to investigate whether genetic variations within *TMEM106B* could influence Parkinson’s disease susceptibility.

**Figure 1.**
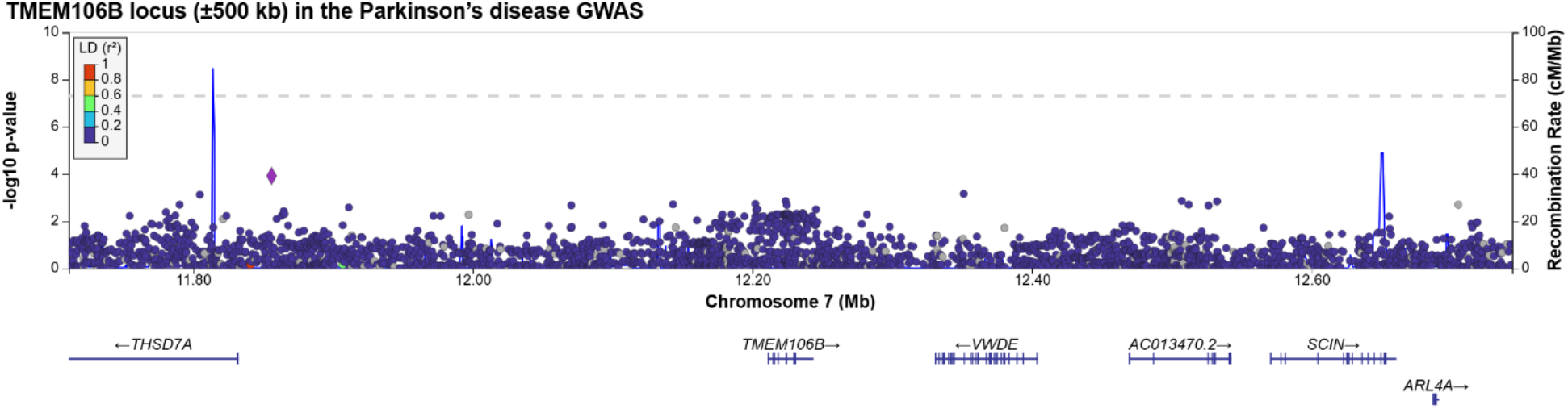
Common variant association signals across the TMEM106B gene region in the largest Parkinson’s disease GWAS. Variants are plotted according to genomic position (GRCh38) and association significance (-log10P). The plot was generated using LocusZoom based on the publicly available GWAS summary statistics (6). No common variants within TMEM106B reached genome-wide significance.

To address this question, we leveraged whole-genome sequencing (WGS) data from the Global Parkinson’s Genetics Program (GP2; https://gp2.org/), the Accelerating Medicines Partnership in Parkinson’s Disease (AMP-PD; https://amp-pd.org/), and the UK Biobank (UKBB; https://www.ukbiobank.ac.uk/). The GP2 cohort consisted of 16,325 cases and 5,561 controls from seven ancestry groups: African (AFR), Admixed American/Latin American (AMR), Central Asian (CAS), East Asian (EAS), European (EUR), Middle Eastern (MDE), and South Asian (SAS). For AMP-PD (*N* cases = 2,241; *N* controls = 3,049) and UKBB (*N* cases = 3,444; *N* controls = 31,940), only European samples were retained for analysis given their limited representation of non-European ancestries. In addition, we performed a meta-analysis across European samples from all three cohorts, yielding a European study population of 15,749 cases and 36,179 controls.

At the sample level, related individuals were excluded, and GP2 samples overlapping with the AMP-PD dataset were removed to prevent sample duplication. At the variant level, we filtered for rare variants (minor allele frequency <0.01) with a minor allele count ≥1, genotype quality ≥25 (when available), heterozygous variant allele fraction between 0.15 and 0.85, and missingness ≤5%. Variant read depth (DP) was filtered using cohort-specific thresholds (DP ≥25 for AMP-PD and UKBB; DP ≥15 for GP2) reflecting differences in variant-calling pipelines, while maintaining comparable genotype quality across cohorts.

Variants were annotated using Ensembl Variant Effect Predictor (VEP) v115 on the GRCh38 reference genome (8), incorporating ClinVar classifications, Combined Annotation Dependent Depletion (CADD) v1.7 scores, AlphaMissense predictions, and gnomAD genome allele frequencies. Gene-based rare-variant association analyses were performed using the Sequence Kernel Association Test-Optimal (SKAT-O, R package) across several variant categories, including all exonic, nonsynonymous, and predicted deleterious variants (CADD ≥20), with subsequent meta-analysis using MetaSKAT (R package). Multiple testing correction was applied using false discovery rate (FDR) adjustment across all variant sets within each ancestry and cohort.

Summary-data-based Mendelian randomization was performed using software from Yang Lab (9) to assess whether genetically regulated TMEM106B expression is causally associated with Parkinson’s disease risk. Significant cis-eQTLs (p ≤ 5×10-6) within cis-window thresholds of ±500 kb were selected from the MetaBrain (10) cortical tissue dataset (n = 2865) for this analysis.

A total of 4,581 unique rare variants were identified across all three cohorts. In the GP2 cohort, 33 nonsynonymous and two loss-of-function (LoF) variants were identified, with the distribution across ancestry groups detailed in Table 1. The largest numbers of nonsynonymous variants were observed in the EAS and EUR groups, consistent with their larger sample sizes. In the AMP-PD cohort, nine nonsynonymous variants were identified, whereas no LoF variants were observed. In the UKBB cohort, 29 nonsynonymous variants and two LoF variants were identified. No pathogenic or likely pathogenic variants according to ClinVar were observed in any cohort.

**Table 1.** Rare *TMEM106B* variant counts and rare variant SKAT-O test results across cohorts.

| Cohort |  | Cases (N) | Controls (N) | Variants tested (N) | Variant consequence |  | SKAT-O p-values* |  |  |
| --- | --- | --- | --- | --- | --- | --- | --- | --- | --- |
| | | | | | NS | LoF | NS | CADD $\geq 20$ | LoF |
| GP2 | AFR | 1,370 | 2,029 | 287 | 2 | 0 | 0.644 | 0.299 | - |
|  | AMR | 755 | 287 | 381 | 5 | 0 | 0.160 | 0.241 | - |
|  | CAS | 330 | 340 | 165 | 3 | 0 | 0.705 | 0.752 | - |
|  | EAS | 2,617 | 1,026 | 642 | 8 | 1 | 0.299 | 0.061 | 0.065 |
|  | EUR | 10,064 | 1,190 | 1257 | 7 | 1 | 0.602 | 0.519 | 0.566 |
|  | MDE | 738 | 405 | 247 | 2 | 0 | 0.247 | 0.247 | - |
|  | SAS | 451 | 284 | 255 | 6 | 0 | 0.548 | 0.288 | - |
| AMP-PD |  | 2,241 | 3,049 | 1,163 | 9 | 0 | 0.257 | 0.350 | - |
| UKBB |  | 3,444 | 31,940 | 1,977 | 29 | 2 | 0.607 | 0.440 | 0.257 |
| EUR Meta-analysis |  | 15,749 | 36,179 | - | - | - | 0.825 | 0.424 | 0.404 |

AFR: African; AMR: Admixed American/Latin American; CAS: Central Asian; EAS: East Asian; EUR: European; MDE: Middle Eastern; SAS: South Asian; AMP-PD: Accelerating Medicines Partnership in Parkinson's Disease; UKBB: UK Biobank; NS: nonsynonymous; LoF: loss-of-function; CADD: Combined Annotation Dependent Depletion; SKAT-O: Sequence Kernel Association Test-Optimal. \*p-values shown are uncorrected; no association remained significant after false discovery rate correction.

Single-variant association analyses revealed no significant enrichment of nonsynonymous or LoF variants in Parkinson’s disease in any cohort or ancestry group. Gene-based burden analyses using SKAT-O similarly identified no significant cumulative effect of *TMEM106B* rare variants on Parkinson’s disease risk in any variant category (nonsynonymous, LoF, or CADD-deleterious) across any cohort or ancestry group (Table 1). Meta-analysis of EUR ancestry samples across all three cohorts confirmed the absence of a significant burden signal (Table 1). These results are in line with previous large-scale rare-variant burden testing efforts in Parkinson’s disease, which similarly did not identify *TMEM106B* as a significant contributor to disease risk (11).

Summary-data-based Mendelian randomization analysis using MetaBrain eQTL data did not identify a significant pleiotropic association between *TMEM106B* expression and Parkinson’s risk (beta = −0.068, p = 0.055; HEIDI p = 0.092). However, the direction of effect was consistent with the findings of Meng et al., in which TMEM106B deficiency exacerbated α-synuclein pathology (1).

Pathogenic variants in *TMEM106B* have been associated with hypomyelinating leukodystrophy 16 (OMIM #617964), an autosomal dominant neurological disorder of infancy characterized by hypotonia, nystagmus and neurodevelopmental delay (12). The autosomal dominant inheritance pattern of this condition, together with apparent high penetrance of pathogenic variants, supports the expectation that such variants would be extremely rare or absent in the general population and unlikely to be associated with later-onset phenotypes such as Parkinson’s disease. Consistent with this hypothesis, we identified no pathogenic or likely pathogenic variants in any cohort, and observed no significant associations between nonsynonymous or LoF variants and Parkinson’s disease risk in either single-variant or burden analyses.

We note that the relationship between *TMEM106B* and Parkinson’s disease may be more complex than a simple LoF model would suggest. A recent study reported that TMEM106B knockdown had neuroprotective effects in PD models via regulation of the autophagy-lysosome pathway (13), which contrasts with the findings of Meng et al. who showed exacerbated pathology upon TMEM106B deficiency. This discrepancy highlights the complexity of TMEM106B biology and suggests that its effects may be context-dependent, varying with cell type, model system, or disease stage.

In conclusion, using large and ancestry-diverse case-control cohorts, we found no evidence that rare coding variants in *TMEM106B* are associated with PD risk, either individually or through cumulative burden. These results suggest that rare variants in *TMEM106B* do not play a major role in PD susceptibility across the populations studied.

## Data availability statement

Data used in the preparation of this article were obtained from the Global Parkinson’s Genetics Program (GP2; https://gp2.org). Specifically, we used Tier 2 data from GP2 release 11 (10.5281/zenodo.17753486). Tier 2 data access requires approval and the submission of a Data Use Agreement signed by your institution. Qualified researchers are encouraged to apply for direct access to the data through Accelerating Medicines Partnership in Parkinson’s Disease (AMP®-PD) website (https://amp-pd.org/register-for-amp-pd). This project was completed under UK Biobank application 45551.

## Code Availability

All code generated for this article, and the identifiers for all software programs and packages used, are available on GitHub at https://github.com/GP2code/GP2-TMEM106B-PD and were given a persistent identifier via Zenodo (DOI: 10.5281/zenodo.22088326).

## Acknowledgements

We thank the patients and their families for participating in this study. H.M.C.S. is supported by Fulbright Colombia, the Ministry of Science, Technology and Innovation of Colombia, and the American Parkinson Disease Association. K.S. is supported by Parkinson’s Canada. Z.G.O. is supported by the Fonds de recherche du Québec—Santé (FRQS) Chercheurs-boursiers award, in collaboration with Parkinson Quebec, and is a William Dawson Scholar.

Data used in the preparation of this article were obtained from the Accelerating Medicine Partnership® (AMP®) Parkinson’s Disease (AMP PD) Knowledge Platform. For up-to-date information on the study, visit https://www.amp-pdrd.org. The AMP® PD program is a public-private partnership managed by the Foundation for the National Institutes of Health and funded by the National Institute of Neurological Disorders and Stroke (NINDS) in partnership with the Aligning Science Across Parkinson’s (ASAP) initiative; Celgene Corporation, a subsidiary of Bristol-Myers Squibb Company; GlaxoSmithKline plc (GSK); The Michael J. Fox Foundation for Parkinson’s Research; Pfizer Inc.; AbbVie Inc.; Sanofi US Services Inc.; and Verily Life Sciences. ACCELERATING MEDICINES PARTNERSHIP and AMP are registered service marks of theU.S. Department of Health and Human Services. Clinical data and biosamples used in preparation of this article were obtained from the (i) Michael J. Fox Foundation for Parkinson’s Research (MJFF) and National Institutes of Neurological Disorders and Stroke (NINDS) BioFIND study,(ii) Harvard Biomarkers Study (HBS) and the Stephen & Denise Adams Center for Parkinson’s Disease Research of Yale School of Medicine (CPDR-Y), (iii) National Institute on Aging (NIA) International Lewy Body Dementia Genetics Consortium Genome Sequencing in Lewy Body Dementia Case-control Cohort (LBD), (iv) NINDS Parkinson’s Disease Biomarkers Program (PDBP), and (v) MJFF Parkinson’s Progression Markers Initiative (PPMI). BioFIND is sponsored by The Michael J. Fox Foundation for Parkinson’s Research (MJFF) with support from the National Institute for Neurological Disorders and Stroke (NINDS). The BioFIND Investigators have not participated in reviewing the data analysis or content of the manuscript. Genome sequence data for the Lewy body dementia case-control cohort were generated at the Intramural Research Program of the U.S. National Institutes of Health. The study was supported in part by the National Institute on Aging (program #: 1ZIAAG000935) and the National Institute of Neurological Disorders and Stroke (program #: 1ZIANS003154). The Harvard Biomarker Study (HBS) is a collaboration of HBS investigators and funded through philanthropy and NIH and Non-NIH funding sources. The Stephen & Denise Adams Center for Parkinson’s Disease Research of Yale School of Medicine is funded through philanthropy and NIH and non-NIH funding sources. The HBS and CPDR-Y Investigators have not participated in reviewing the data analysis or content of the manuscript. PPMI is sponsored by The Michael J. Fox Foundation for Parkinson’s Research and supported by a consortium of scientific partners. The PPMI investigators have not participated in reviewing the data analysis or content of the manuscript. The Parkinson’s Disease Biomarker Program (PDBP) consortium is supported by the National Institute of Neurological Disorders and Stroke (NINDS) at the National Institutes of Health. The PDBP investigators have not participated in reviewing the data analysis or content of the manuscript.

This research used the NeuroHub infrastructure and was undertaken thanks in part to funding from the Canada First Research Excellence Fund, awarded through the Healthy Brains, Healthy Lives initiative at McGill University. And our sincere thanks to the participants and researchers of the UK Biobank, who made this effort possible. This project was completed under UK Biobank application 45551. This research was enabled in part by support provided by Calcul Québec and the Digital Research Alliance of Canada.

This project was supported by the Global Parkinson’s Genetics Program (GP2; https://gp2.org), a research program of Aligning Science Across Parkinson’s (ASAP) (https://ror.org/03zj4c476). ASAP is managed by the Coalition for Aligning Science (CAS) (https://ror.org/00rf4ez40). The Michael J. Fox Foundation for Parkinson’s Research (MJFF) (https://ror.org/03arq3225) is the funding and strategic partner. For a complete list of GP2 members see https://doi.org/10.5281/zenodo.7904831.

